# Modulation of motor cortical excitability by continuous theta-burst stimulation in adults with autism spectrum disorder: The roles of *BDNF* and *APOE* polymorphisms

**DOI:** 10.1101/2020.05.11.20083162

**Authors:** Ali Jannati, Mary A. Ryan, Gabrielle Block, Fae B. Kayarian, Lindsay M. Oberman, Alexander Rotenberg, Alvaro Pascual-Leone

## Abstract

**Objective:** To assess the utility of the modulation of motor cortex (M1) excitability by continuous theta-burst stimulation (cTBS) as a physiologic biomarker for adults with autism spectrum disorder (ASD), and to evaluate the influences of brain-derived neurotrophic factor (*BDNF*) and apolipoprotein E (*APOE*) polymorphisms on cTBS aftereffects.

**Methods:** 44 neurotypical individuals (NT; age 21–65, 34 males) and 19 age-matched adults with high-functioning ASD (age 21–58, 17 males) underwent M1 cTBS. Cortico-motor reactivity was assessed before cTBS and thereafter every 5–10 minutes for 60 minutes (T5–T60).

**Results:** Logistic regressions found cTBS-induced change in amplitude of motor evoked potentials (∆MEP) at T15 was a significant predictor of ASD diagnosis (*p*=0.04). ∆MEP at T15 remained a significant predictor of diagnosis among *BDNF* Met+ subjects and *APOE* ε4− subjects (*p*’s < 0.05) but not *BDNF* Met− subjects. ∆MEP at T30 was the best predictor of diagnosis among *APOE* ε4+ subjects (*p* = 0.08).

**Conclusions:** We confirm previous findings on the utility of cTBS measures of plasticity for adults with ASD, and we find the diagnostic utility of cTBS is modulated by *BDNF* and *APOE* SNPs.

**Significance:** It is important to control for *BDNF* and *APOE* polymorphisms when comparing TBS aftereffects in ASD and NT individuals.

## 1. Introduction

Autism spectrum disorder (ASD) refers to a group of complex neurodevelopmental disorders characterized by: (1) persistent deficiencies in social communication and social interaction, and (2) limited interests and repetitive behavior (American Psychiatric Association 2013). ASD has an estimated prevalence of 14.5 per 1,000 in the U.S. (Christensen et al. 2018) and often results in significant impairments in activities of daily living, both in children and adults (Haertl et al. 2013). Because of the large heterogeneity of the clinical endophenotype in ASD and symptom manifestation over a range of ages and to different degrees, the clinical diagnosis of ASD can be challenging and is typically based on behavioral interviews and subjective clinical impression. Thus, an objective neurophysiologic biomarker that can facilitate ASD diagnosis is highly desirable, especially to improve diagnostic accuracy and to enable metrics of target engagement and clinical/behavioral outcomes in therapeutic interventions.

Data from rodent models of ASD and studies on monogenic causes of ASD in humans indicate aberrant synaptic plasticity mechanisms in the pathophysiology of ASD, including use-dependent changes in synaptic strength (Bourgeron 2009; Krueger and Bear 2011; Percy 2011; Pega et al. 2011; Bhakar et al. 2012; Gipson and Johnston 2012). Animal studies have found abnormalities in long-term potentiation (LTP) and long-term depression (LTD) mechanisms of synaptic plasticity in ASD (Huber et al. 2002; Narita et al. 2002; Dani et al. 2005; Rinaldi et al. 2007, 2008a, 2008b; Tordjman et al. 2007; Gogolla et al. 2009).

Plasticity mechanisms similar to LTP and LTD can be studied noninvasively in humans using transcranial magnetic stimulation (TMS) (Barker et al. 1985; Ziemann 2004; Thickbroom 2007; Barker 2017). TMS is a neurophysiological technique based on the principle of electromagnetic induction that enables triggering or modulation of neural activity in the brain (Hallett 2007) and is considered safe when applied following the recommended guidelines (Rossi et al. 2009; Rossini et al. 2015). Delivering a single TMS pulse (spTMS) to primary motor cortex (M1) can induce a motor evoked potential (MEP) in the target muscle. TMS has been used in various forms including spTMS, paired-pulse TMS (ppTMS), and repetitive TMS (rTMS) at specific intensities, frequencies, and patterns of stimulation to study, modify, or restore activity in corticospinal pathways, as well as motor and non-motor brain regions and networks (see Valero-Cabré et al. 2017 for a review).

Several spTMS studies in ASD have found no significant difference in baseline M1 excitability in ASD (Théoret et al. 2005; Minio-Paluello et al. 2009; Enticott et al. 2012, 2013b). PpTMS studies have found no alteration in short-interval intracortical inhibition (SICI) (Théoret et al. 2005; Jung et al. 2013) or intracortical facilitation (ICF) in ASD (Théoret et al. 2005; Enticott et al. 2010, 2013b). The findings of intracortical inhibition in ASD have been mixed, with some ASD individuals exhibiting abnormal intracortical inhibition while others have responses similar to those of NT individuals (Enticott et al. 2010, 2013b; Oberman et al. 2010).

A form of rTMS referred to as theta-burst stimulation (TBS) of M1 (Huang et al. 2005) consists of 50Hz bursts of triplet TMS pulses repeated at 5 Hz for a total of 600 pulses, in one of two protocols: (1) intermittent theta-burst stimulation (iTBS) with a 2-sec on, 8-sec off pattern for 190 sec that typically induces MEP facilitation by ~35% for up to 60 min; (2) continuous theta-burst stimulation (cTBS) for 40 sec that typically induces MEP suppression by ~25% for up to 50 min (Wischnewski and Schutter 2015). Facilitation and suppression of MEPs by cTBS and iTBS protocols are considered to involve mechanisms similar to LTP and LTD, respectively (Huang et al. 2005; Huerta and Volpe 2009). The return of post-TBS MEP amplitudes to their baseline levels is considered a neurophysiologic index of the efficacy of the mechanisms of cortical plasticity (Pascual-Leone et al. 2005, 2011; Oberman et al. 2010, 2012, 2014, 2016; Tremblay et al. 2015; Suppa et al. 2016). TBS aftereffects involve mechanisms of gamma-aminobutyric acid-(GABA-) ergic and glutamatergic plasticity (Huang et al. 2007, 2008; Stagg et al. 2009; Trippe et al. 2009; Benali et al. 2011).

Studies by Oberman and colleagues (Oberman et al. 2012, 2016) found greater and longer-lasting TBS-induced changes in MEP amplitude in adults with ASD compared to neurotypical (NT) adults, indicating an exaggerated, *hyperplastic* response to TBS in ASD. Recently, we found that children and adolescents with high-functioning (HF) ASD had abnormally greater facilitatory responses to cTBS relative to typically developing children (Jannati et al. 2020). Moreover, cTBS measures of plasticity showed a maturational trajectory in children and adolescents with high-functioning ASD (HF-ASD), in which the extent of, or the maximum, cTBS-induced suppression of MEPs increased linearly with age (Oberman et al. 2014; Jannati et al. 2020).

These results collectively support the utility of cTBS measures of cortical plasticity as potential biomarkers for individuals with ASD across the lifespan. In recent years, however, several studies have documented large inter- and intra-individual variability in M1 cTBS responses among healthy adults (Corp et al. In Press; Hamada et al. 2013; Goldsworthy et al. 2014; Nettekoven et al. 2015; Vallence et al. 2015; Hordacre et al. 2017; Jannati et al. 2017, 2019). Such variability could limit the biomarker utility of cTBS for differentiating individuals with ASD from their NT counterparts. Careful consideration of possible sources of within- and across-individual variability is thus important.

To this end, two single-nucleotide polymorphism (SNPs) have been identified as important contributors to variability of rTMS and other measures of neuroplasticity: the Val66Met SNP in the brain-derived neurotrophic factor (*BDNF*) gene (Cheeran et al. 2008; Antal et al. 2010; Lee et al. 2013; Chang et al. 2014; Di Lazzaro et al. 2015; Fried et al. 2017; Jannati et al. 2017, 2019) and the presence of ε4 allele in the apolipoprotein E (*APOE*) gene (Mahley and Rall Jr 2000; White et al. 2001; Nichol et al. 2009; Wolk et al. 2010; Peña-Gomez et al. 2012; Jannati et al. 2019). Here we address these issues by including a relatively large (n = 44) sample of NT participants and by controlling for *BDNF* or *APOE* SNP when comparing cTBS measures of plasticity between ASD and NT participants. We then calculate the standard measures of biomarker utility for each comparison of cTBS responses.

## 2. Methods

### 2.1. Participants

63 adults (age range 21–65, 12 females) participated in this study, which was approved by the Institutional Review Board at Beth Israel Deaconess Medical Center (BIDMC) in accordance with the Declaration of Helsinki. All participants provided written informed consent prior to enrollment and received monetary compensation upon completion. None of the participants had any TMS contraindication (Rossi et al. 2009), and all had a normal neurological examination. There were two groups: (1) high-functioning adults with non-syndromic ASD (n = 19); (2) neurotypical (NT) age- and gender-matched control participants (n = 44). Participants were recruited through local community advertisements, and local autism associations and clinics. All participants in the ASD group had a documented clinical diagnosis made by a psychiatrist or clinical psychologist, met the Diagnostic and Statistical Manual of Mental Disorders, 5th edition (DSM-5) (American Psychiatric Association, 2013) criteria for ASD, and were independently assessed with the Autism Diagnostic Observation Schedule (ADOS; mean score = 8.68; SD = 4.35). Participants in the NT group had no neurological or psychological disorder. All participants were screened following the safety recommendations endorsed by the International Federation of Clinical Neurophysiology (Rossi et al. 2009). Detailed demographic characteristics of the participants are presented in Table 1 and comparisons of those characteristics between ASD and NT groups are presented in Table 2.

**Table 1.** Participants’ demographics, neuropsychological measures, and single-nucleotide polymor

| Group | Participant ID | Age | Sex | Race | Education* (yr) | Handedness | *BDNF*<sup>†</sup> | *APOE*<sup>‡</sup> | IQ | Verbal KN | Nonverbal FR | ADOS | AQ | Neuroactive medication(s) | Neurological / Psychiatric Comorbidities |
| --- | --- | --- | --- | --- | --- | --- | --- | --- | --- | --- | --- | --- | --- | --- | --- |
| ASD<br>(*n* = 19) |  |  |  |  |  |  |  |  |  |  |  |  |  |  |  |
|  | AS1 | 25 | M | White | 12 | R | Val/Val | ε3/ε3 | 79 | 8 | 5 | 8 | – | Adderall | ADHD |
|  | AS2 | 49 | M | White | 19 | R | Val/Val | ε3/ε4 | 112 | 17 | 7 | 12 | – | Ablify, Ambien, Melatonin, Vyvanse | Anxiety, Depression |
|  | AS3 | 21 | M | Other | – | L | Val/Val | ε3/ε4 | 94 | 9 | 9 | 7 | – | Risperidone, Ritalin | – |
|  | AS4 | 58 | F | White | 16 | R | Val/Val | – | 121 | 18 | 9 | 10 | 36 | Desipramine, Frova, Modafinil | Depression, Migraines |
|  | AS5 | 42 | M | White | 15 | R | Val/Met | ε3/ε3 | 124 | 14 | 14 | 3 | 28 | – | Anxiety, Depression |
|  | AS6 | 33 | F | White | 20 | R | Val/Val | ε3/ε4 | 118 | 17 | 9 | 5 | 35 | Imitrex | Depression, Migraines |
|  | AS7 | 32 | M | White | 13 | R | – | – | 70 | 9 | 1 | 4 | 16 | Ablify, Hydroxyzine, Luvox, Methimazole | Anxiety, Depression, Hyperthyroidism |
|  | AS8 | 27 | M | White | 15 | R | Val/Met | ε3/ε3 | 94 | 8 | 6 | 10 | 33 | Risperidone, Wellbutrin | Anxiety, Depression |
|  | AS9 | 50 | M | White | 12 | R | Val/Met | ε3/ε3 | 103 | 12 | 9 | 8 | 34 | Celexa | Anxiety, Depression, Fainting/Dizzy spells |
|  | AS10 | 52 | M | White | – | L | Val/Val | ε3/ε4 | 118 | 19 | 7 | 3 | 43 | – | ADHD, Anxiety |
|  | AS11 | 52 | M | White | 11 | R | – | – | 100 | 9 | 11 | 5 | 36 | – | Anxiety, Migraine |
|  | AS12 | 37 | M | Asian | 18 | R | Val/Met | ε3/ε3 | 115 | 15 | 10 | 16 | 21 | – | ADHD, Depression |
|  | AS13 | 28 | M | White | 20+ | R | Val/Met | ε3/ε4 | 121 | 16 | 11 | 7 | 20 | – | Dysthymia |
|  | AS14 | 50 | M | – | 16 | R | Val/Val | ε3/ε3 | 112 | 12 | 12 | 10 | 32 | Zoloft | Social Anxiety Disorder |
|  | AS15 | 21 | M | Asian | 13 | R | Val/Met | ε3/ε3 | 124 | 16 | 12 | 15 | 26 | Lexapro | Anxiety, Depression |
|  | AS16 | 34 | M | White | 19 | R | Val/Met | ε3/ε4 | 118 | 13 | 13 | 4 | 35 | – | – |
|  | AS17 | 31 | M | White | – | R | Val/Val | ε3/ε3 | 79 | 9 | 4 | 17 | – | Buspar, Lexapro | Anxiety, Depression |
|  | AS18 | 41 | M | White | 17 | R | Val/Met | ε3/ε3 | 109 | 10 | 13 | 13 | 26 | Acetyl-L-Carnitine | Depression, Essential tremor |
|  | AS19 | 41 | M | White | 16 | R | Val/Met | ε3/ε3 | 88 | 4 | 12 | 8 | 17 | – | – |
| NT<br>(*n* = 44) |  |  |  |  |  |  |  |  |  |  |  |  |  |  |  |
|  | NT1 | 51 | M | White | 16 | R | Val/Val | ε3/ε3 | 115 | 14 | 11 | – | 21 | – | – |
|  | NT2 | 56 | F | Black | 14 | R | Val/Val | ε3/ε4 | 97 | 9 | 10 | – | 9 | – | – |
|  | NT3 | 50 | F | Black | 12 | R | – | – | 97 | 10 | 9 | – | 10 | – | – |
|  | NT4 | 26 | M | Other | 19 | R | Val/Val | ε3/ε4 | 121 | 17 | 10 | – | 23 | – | – |
|  | NT5 | 28 | F | White | – | R | Val/Val | ε3/ε3 | 100 | 10 | 10 | – | 16 | – | – |
|  | NT7 | 46 | M | White | 13 | R | Val/Val | ε3/ε4 | 88 | 9 | 7 | – | 21 | – | – |
|  | NT8 | 28 | M | White | – | R | Val/Val | ε3/ε3 | 115 | 13 | 12 | – | 19 | – | Headaches, Fainting/Dizzy spells |
|  | NT9 | 53 | M | White | 16 | R | Val/Val | ε3/ε4 | 106 | 13 | 9 | – | 11 | – | – |
|  | NT10 | 62 | M | White | 16 | R | Val/Met | ε3/ε4 | 118 | 12 | 14 | – | 25 | – | – |
|  | NT11 | 47 | M | Black | 18 | R | Val/Met | ε3/ε3 | 88 | 9 | 7 | – | 15 | – | – |
|  | NT12 | 32 | M | White | 19 | R | Val/Val | ε3/ε3 | 100 | 9 | 11 | – | 14 | – | Anxiety, Depression |
|  | NT13 | 24 | M | Asian | 16 | R | Val/Met | ε3/ε3 | 100 | 10 | 10 | – | 14 | – | – |
|  | NT14 | 32 | M | Asian | 20+ | R | Val/Met | ε3/ε4 | 103 | 8 | 13 | – | 11 | – | – |
|  | NT15 | 23 | M | Multiracial | 16 | R | Val/Val | ε3/ε3 | 106 | 11 | 11 | – | 20 | – | – |
|  | NT16 | 22 | M | Asian | 17 | R | Val/Met | ε3/ε4 | 103 | 9 | 12 | – | 16 | – | – |
|  | NT17 | 23 | M | White | 17 | R | Val/Val | ε2/ε3 | 127 | 16 | 13 | – | 11 | – | – |
|  | NT18 | 65 | M | Asian | 20+ | R | Val/Val | ε3/ε4 | 124 | 15 | 13 | – | 14 | – | – |
|  | NT19 | 24 | M | Multiracial | 17 | R | Val/Met | ε2/ε3 | 115 | 12 | 13 | – | 12 | – | – |
|  | NT20 | 47 | M | White | 20+ | R | Val/Val | ε3/ε3 | 133 | 17 | 14 | – | 11 | – | Migraine |
|  | NT21 | 22 | M | White | 17 | R | Val/Val | ε2/ε3 | 112 | 10 | 14 | – | 7 | – | – |
|  | NT22 | 21 | M | White | 15 | R | Val/Val | ε3/ε4 | 109 | 14 | 9 | – | 12 | – | – |
|  | NT23 | 21 | M | Multiracial | 16 | R | – | – | 112 | 11 | 13 | – | 7 | – | – |
|  | NT24 | 21 | M | Asian | 16 | R | – | – | 94 | 8 | 10 | – | 17 | – | – |
|  | NT25 | 25 | M | White | 16 | R | Val/Met | ε3/ε3 | 130 | 17 | 13 | – | 16 | – | – |
|  | NT26 | 56 | M | White | 16 | R | Val/Met | ε3/ε3 | 109 | 13 | 10 | – | 16 | – | – |
|  | NT27 | 23 | M | White | 17 | R | Val/Met | ε3/ε3 | 118 | 13 | 13 | – | 22 | – | – |
|  | NT28 | 30 | M | White | 20 | L | Val/Met | ε3/ε4 | 97 | 10 | 9 | – | 13 | – | – |
|  | NT29 | 47 | M | Black | 18 | R | Val/Val | ε3/ε4 | 118 | 16 | 10 | – | 11 | – | – |
|  | NT30 | 45 | M | White | 16 | R | Val/Val | ε2/ε3 | 94 | 9 | 9 | – | 14 | – | – |
|  | NT31 | 23 | M | Black | 14 | R | Val/Val | ε2/ε3 | 103 | 9 | 12 | – | 8 | – | – |
|  | NT32 | 30 | M | White | 18 | R | Val/Met | ε3/ε3 | 109 | 10 | 13 | – | 11 | – | – |
|  | NT33 | 49 | M | Asian | 16 | R | Val/Met | ε3/ε3 | 112 | 12 | 12 | – | 22 | – | – |
|  | NT34 | 28 | M | Multiracial | 18 | R | Val/Val | ε3/ε3 | 100 | 9 | 11 | – | 13 | – | – |
|  | NT35 | 22 | F | White | 16 | R | Val/Val | ε3/ε3 | 100 | 10 | 10 | – | 6 | – | – |
|  | NT36 | 24 | M | White | 13 | R | Val/Met | ε3/ε4 | 118 | 12 | 14 | – | 17 | Cannabis | Anxiety, Depression |
|  | NT37 | 25 | M | Asian | 18 | R | Val/Val | ε3/ε3 | 91 | 7 | 10 | – | 17 | – | – |
|  | NT38 | 23 | M | Asian | 17 | R | Val/Val | ε3/ε3 | 115 | 11 | 14 | – | 19 | – | – |
|  | NT39 | 23 | F | Other | 16 | R | Val/Met | ε3/ε3 | 118 | 14 | 12 | – | 10 | Sumatriptan | Migraine |
|  | NT40 | 25 | F | Asian | 17 | R | Val/Val | ε3/ε4 | 94 | 8 | 10 | – | 28 | – | – |
|  | NT41 | 24 | F | White | 17 | R | Val/Met | ε3/ε3 | 106 | 15 | 7 | – | 14 | – | Fainting/Dizzy spells |
|  | NT42 | 22 | M | White | 16 | R | Val/Val | ε3/ε3 | 118 | 17 | 9 | – | 18 | – | – |
|  | NT43 | 29 | F | White | 18 | R | – | – | 118 | 12 | 14 | – | 9 | – | Hypothyroidism |
|  | NT44 | 25 | F | Other | 20 | R | – | – | 106 | 12 | 10 | – | 10 | – | – |
|  | NT45 | 23 | F | Asian | 16 | L | Met/Met | ε4/ε4 | 103 | 14 | 7 | – | 19 | – | – |

Racial categories were defined according to the National Institutes of Health policy and guidelines on the inclusion of minorities as subjects in clinical research (NIH Office of Extramural Research, 2001). IQ scores were estimated using the Abbreviated Battery of Stanford-Binet IV intelligence scale. *APOE*, apolipoprotein E; AQ, autism-spectrum quotient; ASD, autism spectrum disorder; *BDNF*, brain-derived neurotrophic factor; IQ, intelligence quotient; Met, metionine; NT, neurotypical; Val, Valine. Values marked by "–" were either not reported by the participant or not available.
\*Education data were available for 16 and 42 participants in the ASD and NT groups, respectively.
†*BDNF* data were available for 17 and 40 participants in the ASD and NT groups, respectively.
‡ *APOE* data were available for 16 and 40 participants in the ASD and NT groups, respectively.

### 2.2. Neuropsychological testing

The Abbreviated Battery of Stanford-Binet IV intelligence scale (Thorndike et al. 1986) and the Autism-Spectrum Quotient (AQ) (Baron-Cohen et al. 2001) were completed for both groups. All participants included in the ASD group had an Abbreviated IQ ≥ 70 (Table 1). AQ scores were used to quantify where each participant was situated on the continuum from autism to normality and to rule out clinically significant levels of autistic traits in the NT group (Baron-Cohen et al. 2001). The Autism Diagnostic Observation Schedule (ADOS) Module 4 was conducted to assess the current social-communicative behavior in the ASD group (Lord et al. 2000).

### 2.3. Genetic testing

Saliva samples from 56 participants including 17 participants (89.5%) in the ASD group and 39 participants (88.6%) in the NT group were assessed for single-nucleotide polymorphisms (SNPs) in the brain-derived neurotrophic factor (*BDNF*), Val66Met, and *APOE* genes. Two participants in the ASD group and four participants in the NT group did not consent to providing DNA samples, and one NT sample was deemed unusable.

Aliquot (700 μL) extraction of genomic DNA was performed on saliva samples collected using the Oragene Discover OGR-250 Kit (DNA Genotek Inc., Ottawa, ON, Canada). DNA was extracted from samples using standard methodology and the prepIT•L2P reagent (DNA Genotek Inc. 2015). The following quality control metrics were performed on each sample: PicoGreen fluorometry for double stranded DNA quantification, Nanodrop spectrophotometry as an estimate of sample purity using A260/A280 ratios and agarose gel electrophoresis for visualization of DNA integrity.

The rs6265 SNP of the *BDNF* gene, the rs429358 and the rs7412 SNPs of the *APOE* gene were analyzed using a TaqMan single tube genotyping assay, which uses polymerase chain reaction (PCR) amplification and a pair of fluorescent dye detectors that target the SNP. During PCR, the polymerase releases the fluorescent probe into solution where it is detected using endpoint analysis in an Applied Biosystems, Inc. (Foster City, CA, USA) 7900HT Real-Time instrument.

### 2.4. Transcranial magnetic stimulation

Participants were seated in a comfortable chair with the right arm and hand in a natural pronated ~90° angle on a pillow in front of them. They were instructed to keep their right hand as still and relaxed as possible throughout the experiment. They were also monitored for drowsiness and were asked to keep their eyes open during the TMS application.

All TMS procedures followed the recommended guidelines endorsed by the International Federation of Clinical Neurophysiology (Rossi et al., 2009; Rossini et al., 2015). Single TMS pulses and cTBS were applied to the left primary motor cortex (M1) at 120% of individual resting motor threshold (RMT) and 80% of active motor threshold (AMT), respectively, as biphasic pulses with an antero–posterior–postero-anterior (AP-PA) induced current direction in the brain using a MagPro X100 stimulator and a MC-B70 Butterfly Coil (outer diameter: 97mm; MagPro, MagVenture A/S, Farum, Denmark). The coil was held tangentially to the participant’s head surface, with the handle pointing occipitally and positioned at 45° relative to the mid-sagittal axis of the participant’s head. The optimal spot for the maximal responses of the right first dorsal interosseous (FDI) muscle (“motor hotspot”) was localized. A Polaris infrared-optical tracking system (Northern Digital Inc., Waterloo, ON, Canada) and a frameless stereotactic neuronavigation system (Brainsight, Rogue Research Inc., Montreal, QC, Canada) was used to ensure consistent targeting throughout the experiment. Each participant’s head was registered to the MRI template using defined cranial landmarks and 12 additional points on the scalp to ensure the coil position and orientation were consistent with the MRI (Ruohonen and Karhu 2010).

Surface electromyogram (EMG) was recorded from the right FDI with a PowerLab 4/25 data-acquisition device and LabChart software (AD Instruments, Colorado Springs, CO, USA). Electrodes were placed over the FDI belly (negative) and the first interphalangeal joint of the second finger (positive). The ground electrode was placed over the ipsilateral ulnar styloid process. The TMS system delivered triggered pulses that synchronized the TMS and EMG systems. EMG signal was digitized at 1 kHz for 500 ms following each stimulus trigger and 100 ms pre-trigger, amplified with a range of ±10 mV (band-pass filter 0.3–1000 Hz).

Each TMS session began by localizing the motor hotspot for FDI and assessment of RMT, defined as the lowest intensity of stimulation that elicited motor evoked potentials (MEPs) ≥ 50 μV in at least five of ten pulses in the relaxed right FDI. To assess pre-cTBS cortico-motor reactivity, three blocks of 30 single TMS pulses were applied to M1, with a 5–10 minute inter-block interval and at a random 4–6 s inter-pulse interval. In each block, individual MEPs > 2.5 SD from the mean were excluded. Recent studies have found applying at least 20 single TMS pulses yields a reliable estimate of MEP amplitude at a given time point (Chang et al. 2016; Goldsworthy et al. 2016). Live EMG was monitored to maintain hand relaxation throughout the experiment and to ensure the background EMG activity did not exceed ~100 μV.

Baseline MEP amplitude was calculated as the average of the peak-to-peak amplitude of MEPs in the three blocks. AMT was then assessed as the lowest intensity that elicited MEPs ≥ 200 μV in at least five of ten pulses with the FDI slightly contracted. Live EMG was monitored during the AMT assessment to ensure consistent contraction between ~100–200 μV. After a 5-minute break, during which participants were instructed to maintain hand relaxation to control the effects of voluntary hand movements on cTBS responses (Iezzi et al., 2008), cTBS was applied as 200 bursts of three pulses at 50 Hz, repeated at 200-ms intervals for 40 s (for a total of 600 pulses). Cortico-motor reactivity was reassessed at 5, 10, 15, 20, 30, 40, 50, and 60 min post-cTBS (*T5–T60*).

### 2.5. Statistical analyses

Study data were collected and managed using Research Electronic Data Capture (REDCap) electronic data capture tools hosted at Beth Israel Deaconess Medical Center (Harris et al. 2009, 2019). Stata 14.0 (StataCorp, College Station, TX, USA) and MATLAB R2016b (The MathWorks, Natick, MA, USA) were used for data analyses.

Data from each TMS visit included: (a) RMT and AMT, expressed as percentage of maximum stimulator output (MSO); (b) baseline MEP amplitude, calculated as the average of baseline MEP amplitude in 3 blocks of 30 single TMS pulses, which are expected to provide reliable estimates of MEP amplitude at a given time point (Chang et al. 2016; Goldsworthy et al. 2016); and (c) percent change in the average amplitude of 30 MEPs at T5–T60 relative to baseline (%∆) for each participant. The Shapiro–Wilk found significant deviations in MEP values from normal distribution. Thus, we first baseline-corrected each post-cTBS amplitude by dividing it by the average baseline MEP amplitude in each participant. We then natural log-transformed the baseline-corrected MEP amplitudes at each post-cTBS time point (∆*MEP*) (Nielsen 1996a, 1996b; Pasqualetti and Ferreri 2011). Grand-average ∆MEPs were calculated separately for each time-point in each group. ∆MEP at T5–T20 for one NT subject and ∆MEP at T15 in another NT subject were not obtained due to technical difficulties.

Pairwise comparisons were conducted to assess whether demographic, neuropsychological, genetic, and neurophysiological measures (RMT, AMT, and baseline MEP amplitude) were significantly different between ASD and NT groups (Table 2). Continuous variables were compared using Student’s t-tests, while proportions were compared using Fisher’s Exact tests. All analyses were two-tailed, and α level was set to 0.05.

**Table 2.**
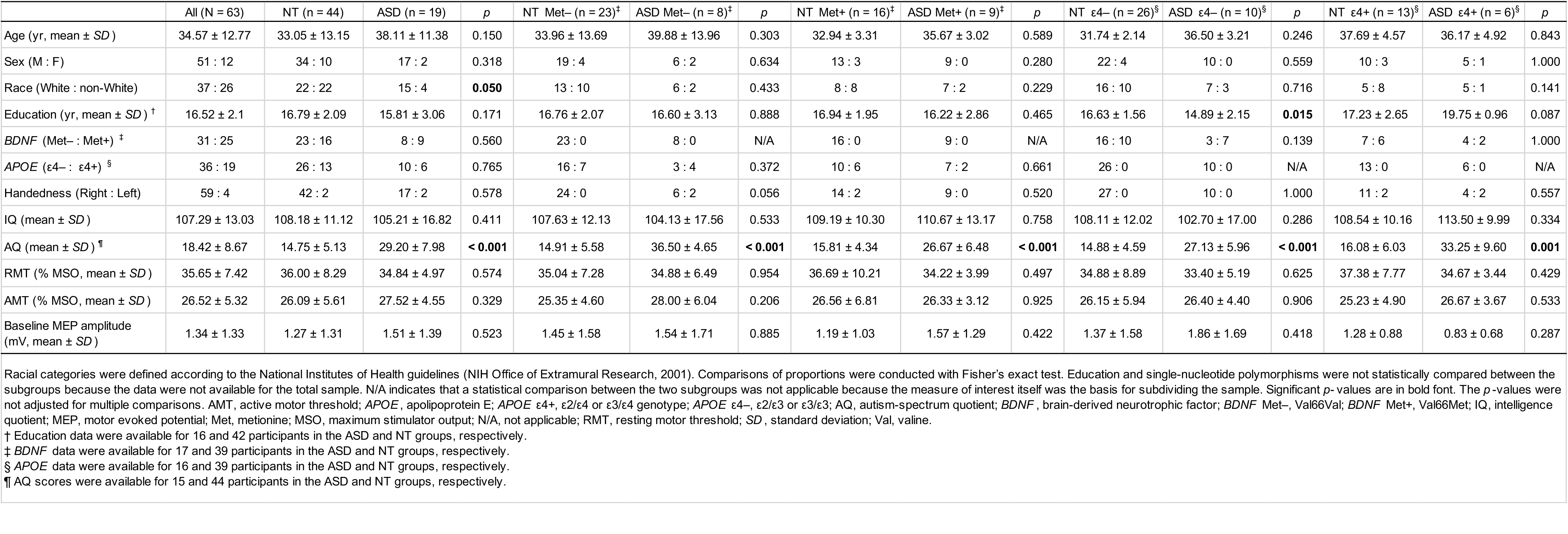
Participants’ demographics, single-nucleotide polymorphisms, neuropsychological scores, and baseline neurophysiological measures.

To compare cTBS aftereffects between ASD and NT groups, ∆MEPs were entered into a 2 (*Diagnosis*) × 8 (*Time*) repeated-measures analysis of variance (ANOVA). However, as the maximum modulation of MEP amplitudes typically occurs within the first 20 minutes after cTBS (Table 4 in Wischnewski and Schutter 2015), planned pairwise comparisons between ∆MEPs at T5–T20 in the two groups were conducted using Student’s t-tests. When indicated, false discovery rate (FDR) was controlled by adjusting the *p*-values for multiple comparisons using the Benjamini-Hochberg method (Benjamini and Hochberg 1995). T15 was selected *post hoc* (see Results) as the time-point at which cTBS plasticity measures were most altered in ASD relative to NT controls.

To assess the predictive utility of cTBS aftereffects, logistic-regression analyses were conducted with *Diagnosis* (ASD vs. NT) as dependent variable (DV) and ∆*MEP* at T15 as the main independent variable (IV). To control for potential confounders, demographic (age, sex, race, and education), neuropsychological (IQ), genetic (*BDNF* and *APOE*), and baseline neurophysiological measures (RMT, AMT, baseline MEP amplitude) were added, one at a time, as covariates to the logistic-regression model. For each logistic-regression model, area under the receiver operating characteristic curve (AUROC), positive and negative predictive values (PPV and NPV) and percentage correctly classified (Dx) were calculated.

To control for influences of *BDNF* and *APOE* genotypes on cTBS aftereffects, ∆MEPs of ASD and NT groups were compared separately for *BDNF* Met−, *BDNF* Met+, *APOE* ε4−, and *APOE* ε4+ participants, and were analyzed similarly as described above. For *APOE* ε4+ participants, T30 was selected *post hoc* (see Results) as the time-point at which cTBS aftereffects were most altered in ASD relative to NT controls. Thus, the logistic-regression analyses to predict *Diagnosis* among *APOE* ε4+ participants were conducted with ∆MEP at T30 as the IV.

## 3. Results

Table 1 details demographics, *BDNF* and *APOE* single-nucleotide polymorphisms, and neuropsychological measures for individual participants. Group means ± SD for those measures and their comparisons in the ASD and NT groups as well as in their *BDNF* and *APOE* subgroups are presented in Table 2.

### 3.1. Demographics and neuropsychogical testing

There was no significant group difference in age, sex, handedness, or education (*p*’s > 0.1), but the proportion of White participants was significantly higher in the ASD group than in the NT group (*p* = 0.050). Therefore, to control for a potential racial confounder, we repeated the main-group logistic regression analysis while restricting the samples to White participants.

In the neuropsychological measures, the IQ scores were comparable between the two groups (*p* = 0.41), indicating that ASD participants did not significantly differ from NT controls in terms of overall cognitive function. As expected, participants with ASD had significantly higher AQ scores than their NT counterparts in the whole sample and in all *BDNF* and *APOE* subgroups (*p*’s ≤ 0.001).

### 3.2. Genetic analyses

Among 56 participants with available *BDNF* results, the proportions of *BDNF* Val/Val and Val/Met genotypes were 55.4% and 44.6%, whereas among 55 participants with available *APOE* results, the proportions of *APOE* ε4− and ε4+ genotypes were 65.5% and 34.6%, respectively. The two groups were comparable in *BDNF* Met−: Met+ and *APOE* ε4−: ε4+ ratios (*p*’s > 0.5).

ASD and NT participants in each *BDNF* and *APOE* subgroup were comparable in terms of demographics and IQ scores (*p*’s > 0.05; Table 2). *APOE* ε4− participants were more educated in the NT group than in the ASD group (*p* = 0.015).

### 3.3. Measures of corticospinal excitability and plasticity

All participants tolerated TMS and cTBS with no complications or unexpected side effects. As detailed in Table 2, RMT, AMT, and baseline MEP amplitudes were comparable between the ASD and NT groups (*p*’s > 0.3), and their *BDNF* and *APOE* subgroups (*p*’s > 0.2). These results indicate the ASD participants did not differ from NT controls in baseline corticospinal excitability.

The repeated-measures ANOVA indicated the ∆MEPs at T5–T60 did not vary significantly by *Diagnosis, F*(1,61) = 0.64, *p* = 0.428, 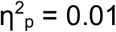, *Time, F*(7,422) = 1.82, *p* = 0.082, 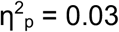, or their interaction, *F*(7,422) = 1.37, *p* = 0.218, 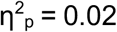. Planned t-tests at T5–T20 showed ASD subjects had significantly greater inhibition of MEPs at T15 (*p* = 0.029). That difference did not remain significant after Bonferroni-Hochberg adjustment (*P_fdr_* = 0.116). ∆MEPs at T5–T10 were not significantly different between the two groups (*p*’s > 0.2). Figure 1 shows the cTBS aftereffects in ASD and NT groups and the ROC curve associated with the logistic regression.

**Figure 1.**
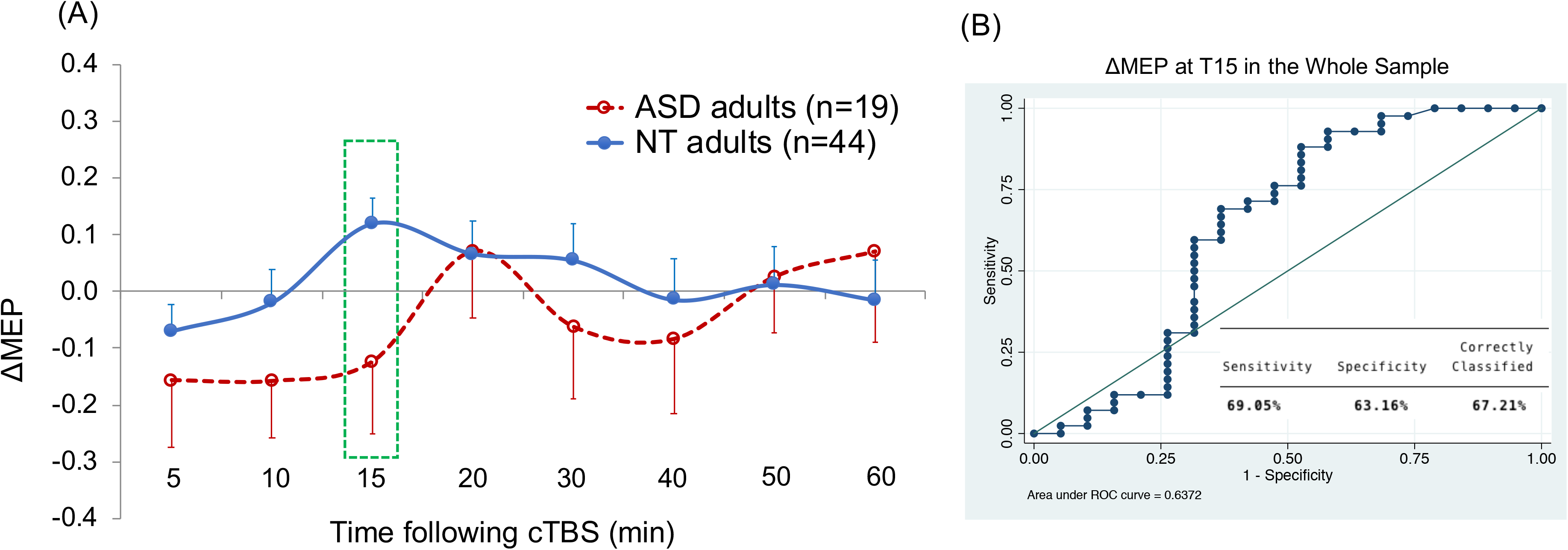
(A) Grand-average ∆MEPs recorded from the right FDI muscle at 5 to 60 minutes following cTBS of the left M1 in ASD and NT groups. ∆MEP at T15 was a significant predictor of diagnosis (*p* = 0.038). Error bars represent standard error of the mean. (B) The ROC curve associated with the logistic regression predicting diagnosis based on T15 ∆MEP. ASD, autism spectrum disorder; cTBS, continuous theta-burst stimulation; ∆MEP, natural log-transformed, baseline-corrected amplitudes of motor evoked potentials; FDI, first dorsal interosseous; M1, motor cortex; NT, neurotypical; ROC, receiver operating characteristic.

Logistic regression analysis found that T15 ∆MEP was a significant predictor of *Diagnosis, β* = −1.55, *z* = 2.07, *p* = 0.038, indicating a more negative ∆MEP at T15 was predictive of ASD diagnosis, AUROC = 0.64, Dx = 67.2%. As detailed in Table 3, follow-up analyses found that T15 ∆MEP remained a significant predictor of *Diagnosis* after controlling for *IQ, Handedness, BDNF, APOE, RMT, AMT*, or *Baseline MEP* (*p*’s < 0.046) but not after controlling for *Age, Sex*, or *Education* (*p*’s > 0.052). However, none of the added covariates was a significant predictor (*p*’s > 0.08), and the predictive powers of all the logistic-regression models were comparable (AUROC range 0.64–0.70). Controlling for *Education* or *Race* resulted in the most predictive models (Dx > 80%), whereas controlling for other covariates resulted in 72–78 % correct classification (Table 3). After limiting the analysis to White participants, T15 ∆MEP remained a significant predictor of *Diagnosis* (*p* = 0.039; AUROC = 0.70; Dx = 75.0%).

**Table 3.**
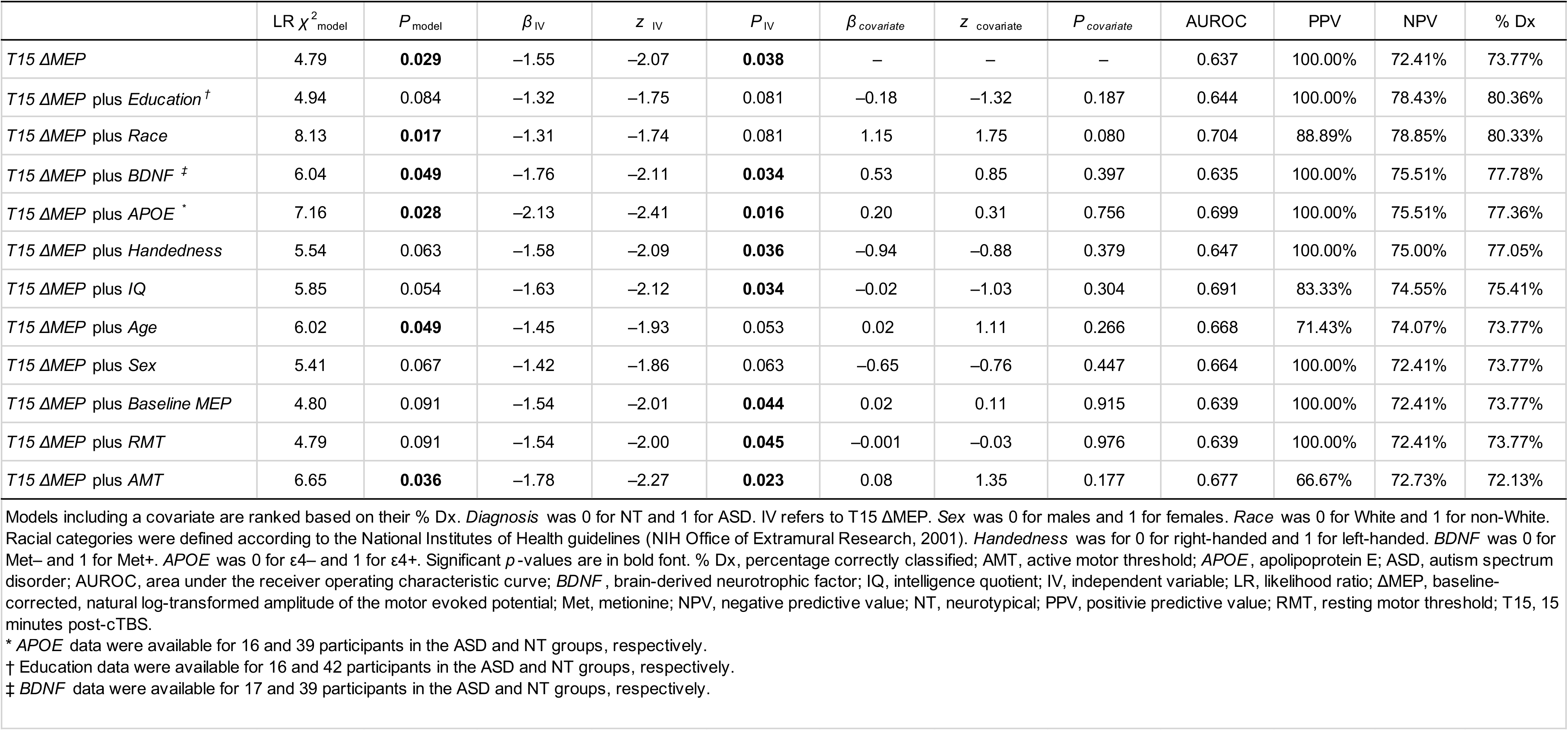
Changes in the logistic regression model predicting diagnosis after adding covariates.

### 3.4. BDNF and APOE influences in cTBS measures of plasticity

Among 53 participants, after controlling for both *BDNF* and *APOE* genotypes, T15 ∆MEP remained a significant predictor of *Diagnosis* (*p* = 0.018; AUROC = 0.69; Dx = 79.3%), whereas neither *BDNF* nor *APOE* status was a significant predictor (*p*’s > 0.2).

Table 4 presents results of the logistic regression analysis predicting *Diagnosis* using T15 ∆MEP as the IV within *BDNF* and *APOE* subgroups. Because controlling for *Education* or *Race* in the overall logistic regression resulted in models with the highest Dx values, *Education* and *Race* were added, one at a time, as covariates to each model. For *APOE* ε4+ participants, T30 ∆MEP was chosen *post hoc* as the time point at which cTBS aftereffects in ASD participants showed the largest alteration compared to NT participants, and similar logistic regression analyses were conducted with T30 ∆MEP as the IV. The analyses found T15 ∆MEP was a significant predictor of *Diagnosis* among 25 *BDNF* Met+ participants (16 NT, 9 ASD; *p* = 0.048; AUROC = 0.69; Dx = 83.3%) and among 36 *APOE* ε4− participants (26 NT, 10 ASD; p = 0.037; AUROC = 0.69; Dx = 79.4%), but not among 31 *BDNF* Met− participants (23 NT, 8 ASD; p = 0.490; AUROC = 0.57; Dx = 73.3%). Figures 2–5 show the comparisons of cTBS aftereffects in *BDNF* and *APOE* subgroups of ASD and NT participants and the ROC curves associated with the logistic regressions (when significant).

**Figure 2.**
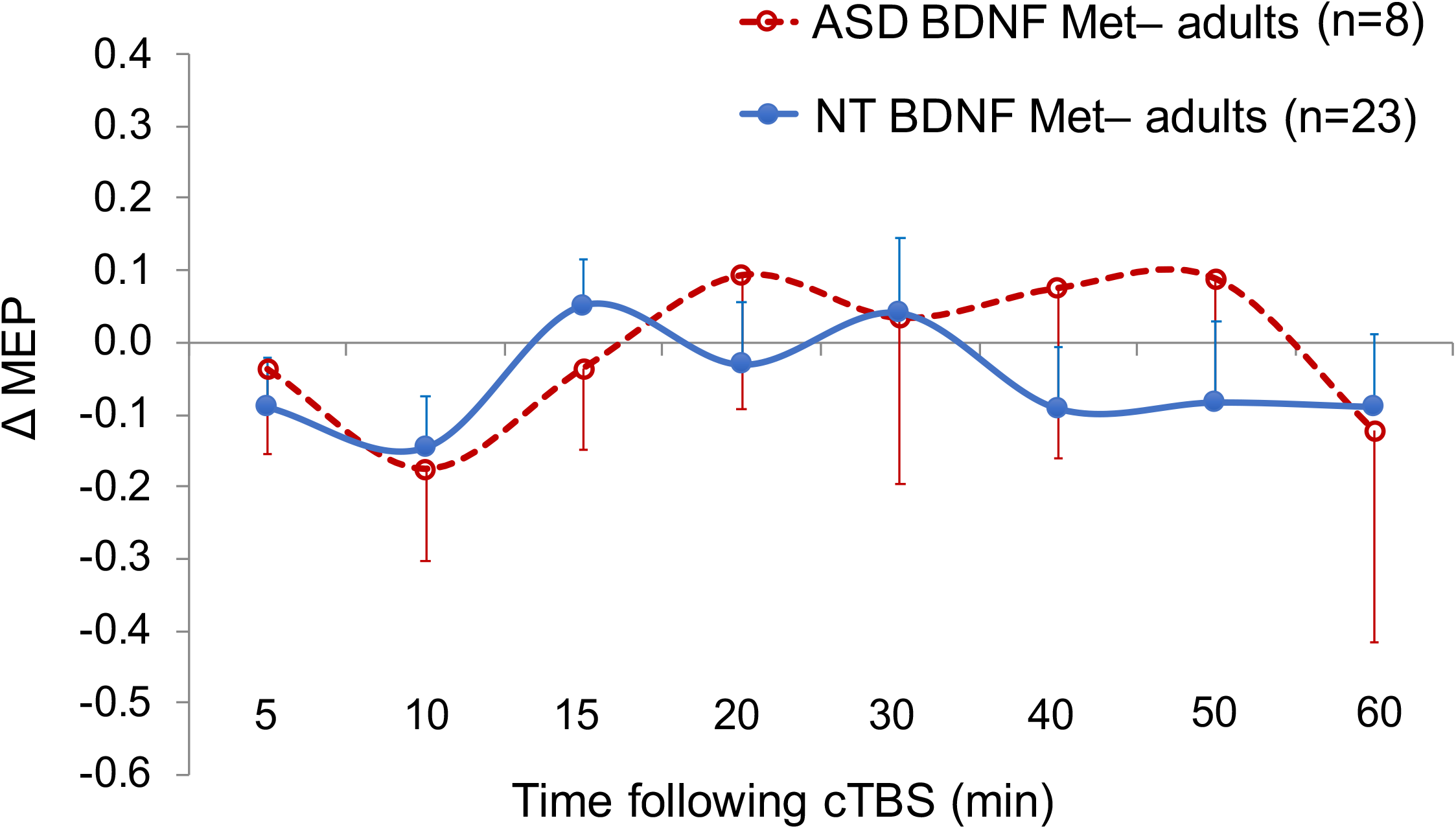
Grand-average ∆MEPs recorded from the right FDI muscle at 5 to 60 minutes following cTBS of the left M1 in *BDNF* Met− subgroups of ASD and NT participants. ∆MEPs were not different between the two groups at any time point. Error bars represent standard error of the mean. ASD, autism spectrum disorder; *BDNF*, brain-derived neurotrophic factor; cTBS, continuous theta-burst stimulation; ∆MEP, natural log-transformed, baseline-corrected amplitudes of motor evoked potentials; FDI, first dorsal interosseous; M1, motor cortex; Met, metionine; NT, neurotypical.

**Figure 3.**
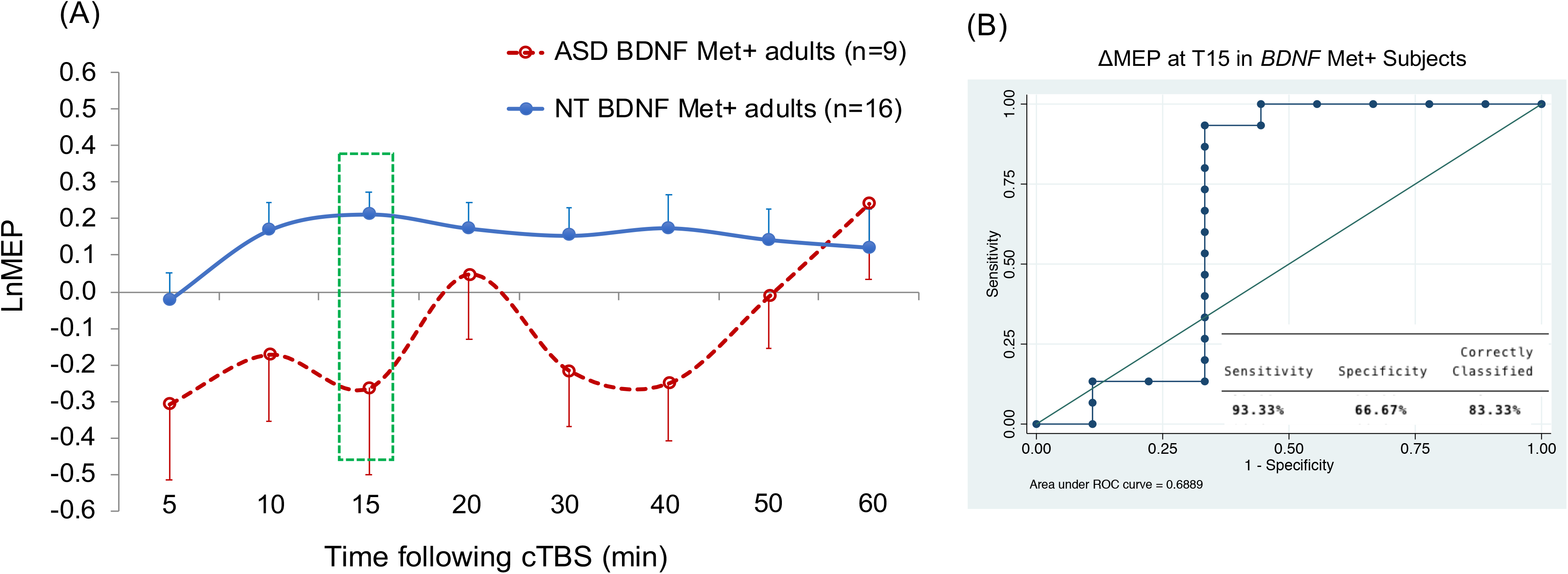
Grand-average ∆MEPs recorded from the right FDI muscle at 5 to 60 minutes following cTBS of the left M1 in *BDNF* Met+ subgroups of ASD and NT participants. ∆MEP at T15 was a significant predictor of diagnosis (*p* = 0.048). Error bars represent standard error of the mean. (B) The ROC curve associated with the logistic regression predicting diagnosis based on T15 ∆MEP. ASD, autism spectrum disorder; *BDNF*, brain-derived neurotrophic factor; cTBS, continuous theta-burst stimulation; ∆MEP, natural log-transformed, baseline-corrected amplitudes of motor evoked potentials; FDI, first dorsal interosseous; M1, motor cortex; Met, metionine; NT, neurotypical; ROC, receiver operating characteristic.

**Figure 4.**
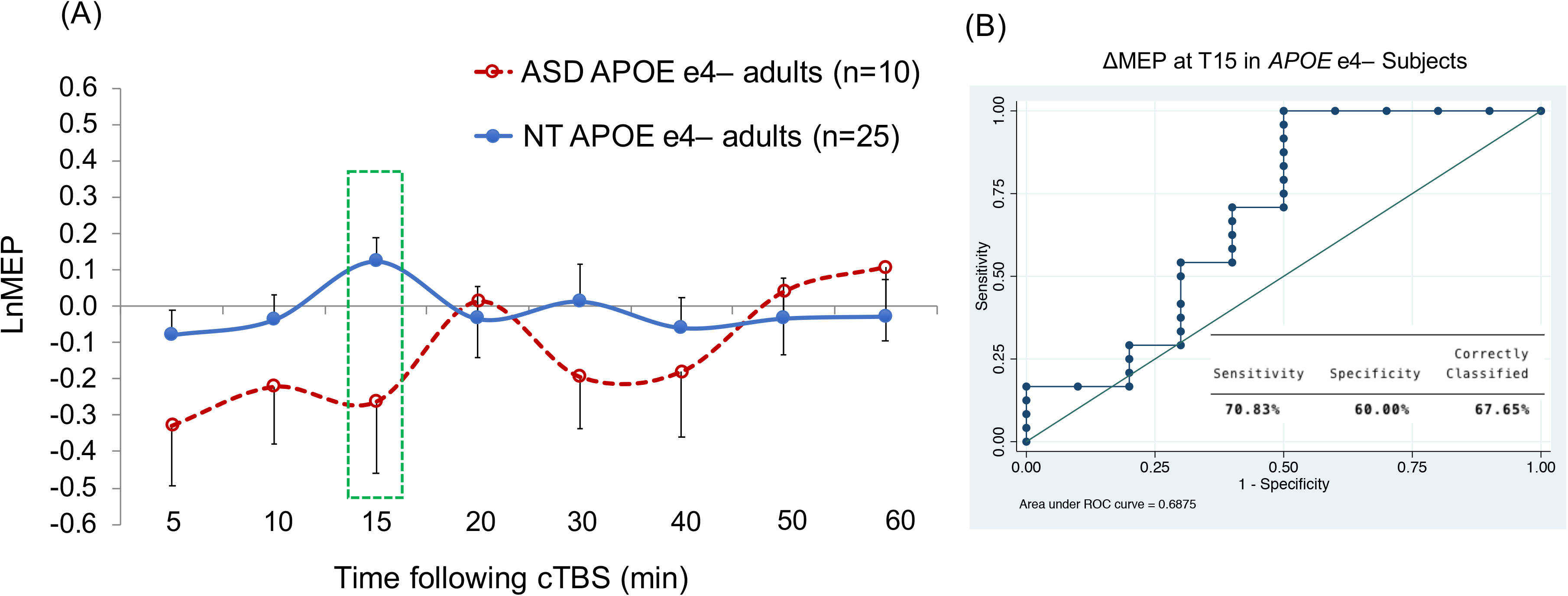
Grand-average ∆MEPs recorded from the right FDI muscle at 5 to 60 minutes following cTBS of the left M1 in *APOE* ε4− subgroups of ASD and NT participants. ∆MEP at T15 was a significant predictor of diagnosis (*p* = 0.037). Error bars represent standard error of the mean. (B) The ROC curve associated with the logistic regression predicting diagnosis based on T15 ∆MEP. *APOE*, apolipoprotein E; ASD, autism spectrum disorder; cTBS, continuous theta-burst stimulation; ∆MEP, natural log-transformed, baseline-corrected amplitudes of motor evoked potentials; FDI, first dorsal interosseous; M1, motor cortex; Met, metionine; NT, neurotypical; ROC, receiver operating characteristic.

**Figure 5.**
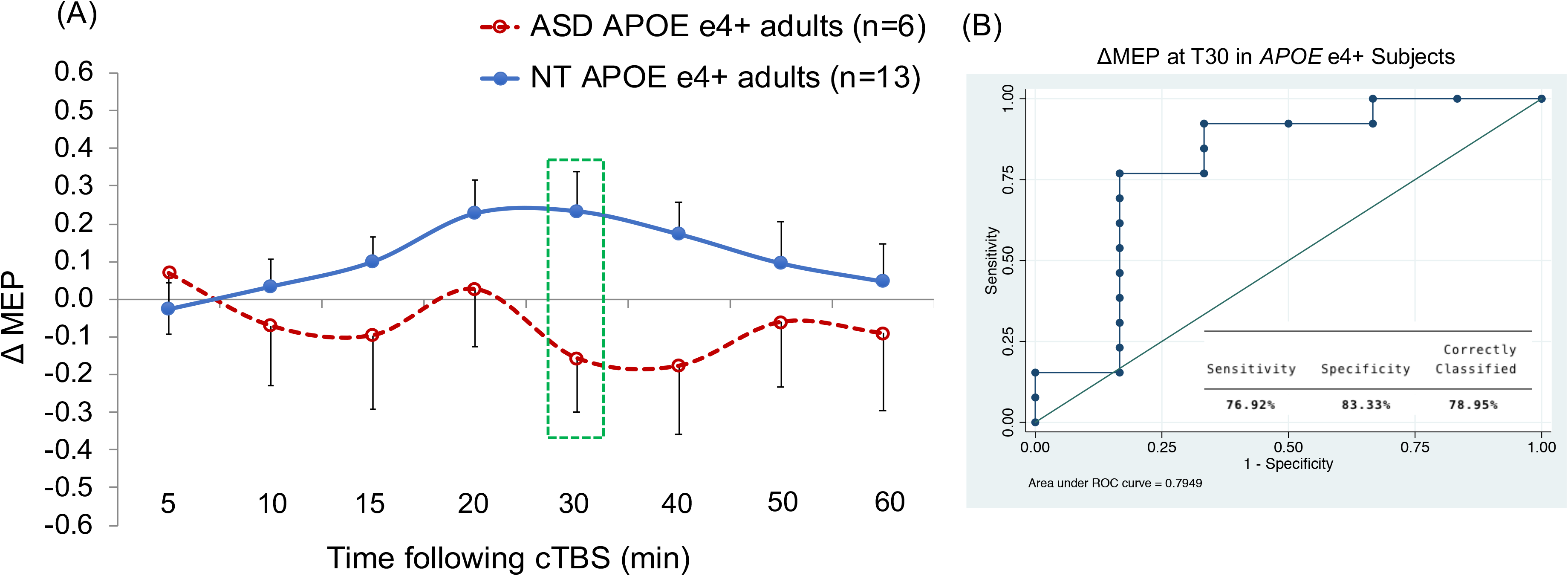
Grand-average ∆MEPs recorded from the right FDI muscle at 5 to 60 minutes following cTBS of the left M1 in *APOE* ε4+ subgroups of ASD and NT participants. The model with ∆MEP at T30 as a predictor was significant (*p* = 0.017). Error bars represent standard error of the mean. (B) The ROC curve associated with the logistic regression predicting diagnosis based on T30 ∆MEP. *APOE*, apolipoprotein E; ASD, autism spectrum disorder; cTBS, continuous theta-burst stimulation; ∆MEP, natural log-transformed, baseline-corrected amplitudes of motor evoked potentials; FDI, first dorsal interosseous; M1, motor cortex; Met, metionine; NT, neurotypical; ROC, receiver operating characteristic.

**Table 4.** The logistic regression models within *BDNF* and *APOE* subgroups.

| | LR $\chi^2_{\text{model}}$ | $P_{\text{model}}$ | $\beta_{\text{IV}}$ | $z_{\text{IV}}$ | $P_{\text{IV}}$ | $\beta_{\text{covariate}}$ | $z_{\text{covariate}}$ | $P_{\text{covariate}}$ | AUROC | PPV | NPV | % Dx |
| --- | --- | --- | --- | --- | --- | --- | --- | --- | --- | --- | --- | --- |
| *BDNF* Met – (n = 31; 23 NT, 8 ASD) |  |  |  |  |  |  |  |  |  |  |  |  |
| *T15 ΔMEP* | 0.50 | 0.480 | –1.0 | –0.69 | 0.490 | – | – | – | 0.571 | – | 73.33% | 73.33% |
| *T15 ΔMEP* plus *Education*<sup>†</sup> | 0.11 | 0.950 | 0.52 | 0.32 | 0.750 | –0.002 | –0.01 | 0.992 | 0.570 | – | 80.00% | 80.00% |
| *T15 ΔMEP* plus *Race* | 1.26 | 0.533 | –1.09 | –0.75 | 0.452 | 0.79 | 0.84 | 0.399 | 0.622 | – | 73.33% | 73.33% |
| *BDNF* Met + (n = 25; 16 NT, 9 ASD) |  |  |  |  |  |  |  |  |  |  |  |  |
| *T15 ΔMEP* | 5.20 | **0.023** | –2.12 | –1.98 | **0.048** | –0.50 | –1.04 | 0.296 | 0.689 | 100.00% | 78.95% | 83.33% |
| *T15 ΔMEP* plus *Education*<sup>†</sup> | 5.25 | 0.073 | –2.09 | –1.92 | 0.055 | –.05 | –.21 | 0.833 | 0.711 | 100.00% | 83.33% | **87.50%** |
| *T15 ΔMEP* plus *Race* | 5.90 | 0.052 | –1.84 | –1.68 | 0.093 | 0.86 | 0.83 | 0.408 | 0.682 | 85.71% | 82.35% | 83.33% |
| *APOE* ε4– (n = 36; 26 NT, 10 ASD) |  |  |  |  |  |  |  |  |  |  |  |  |
| *T15 ΔMEP* | 5.46 | **0.019** | –2.11 | –2.09 | **0.037** | – | – | – | 0.688 | 100.00% | 77.42% | 79.41% |
| *T15 ΔMEP* plus *Education*<sup>†</sup> | 9.48 | **0.009** | –2.03 | –1.85 | 0.064 | –0.63 | –2.04 | **0.041** | 0.828 | 83.33% | 84.00% | **83.87%** |
| *T15 ΔMEP* plus *Race* | 5.48 | 0.065 | –2.15 | –2.06 | 0.040 | –0.12 | –0.14 | 0.892 | 0.671 | 100.00% | 77.42% | 79.41% |
| *APOE* ε4+ (n = 19; 13 NT, 6 ASD) |  |  |  |  |  |  |  |  |  |  |  |  |
| *T15 ΔMEP* | 1.67 | 0.197 | –2.18 | –1.21 | 0.228 | – | – | – | 0.705 | 100.00% | 72.22% | 73.68% |
| *T15 ΔMEP* plus *Education*<sup>†</sup> | 4.42 | 0.110 | –1.68 | –0.86 | 0.388 | 0.63 | 1.51 | 0.131 | 0.789 | – | 73.33% | 64.71% |
| *T15 ΔMEP* plus *Race* | 4.81 | 0.091 | –1.85 | –1.06 | 0.288 | 2.02 | 1.61 | 0.108 | 0.808 | 66.67% | 84.62% | **78.95%** |
| *APOE* ε4+ (n = 19; 13 NT, 6 ASD) |  |  |  |  |  |  |  |  |  |  |  |  |
| *T30 ΔMEP* | 3.69 | 0.055 | –2.63 | –1.62 | 0.106 | – | – | – | 0.795 | 100.00% | 76.47% | 78.95% |
| *T30 ΔMEP* plus *Education*<sup>†</sup> | 5.63 | 0.060 | –1.92 | –1.30 | 0.193 | 0.63 | 1.40 | 0.163 | 0.827 | 50.00% | 84.62% | 76.47% |
| *T30 ΔMEP* plus *Race* | 8.19 | 0.017 | –2.93 | –1.73 | 0.083 | 2.60 | 1.87 | 0.062 | 0.859 | 80.00% | 1.20% | **84.21%** |

Models in each genetic subgroup are ranked based on their % Dx. *Diagnosis* was 0 for NT and 1 for ASD. IV refers to ΔMEP at T15 or T30, as specified. *Race* was 0 for White and 1 for non-White. Racial categories were defined according to the National Institutes of Health guidelines (NIH Office of Extramural Research, 2001). Significant *p*-values are in bold font. In each subgroup, the highest % Dx value of a significant model is in bold font. % Dx, percentage correctly classified; *APOE*, apolipoprotein E; ASD, autism spectrum disorder; AUROC, area under the receiver operating characteristic curve; *BDNF*, brain-derived neurotrophic factor; IV, independent variable; LR, likelihood ratio; ΔMEP, baseline-corrected, natural log-transformed amplitude of the motor evoked potential; Met, metionine; NPV, negative predictive value; NT, neurotypical; PPV, positive predictive value; *Tn*, *n* minutes post-cTBS.
\* *APOE* data were available for 16 and 39 participants in the ASD and NT groups, respectively.
† Education data were available for 16 and 42 participants in the ASD and NT groups, respectively.
‡ *BDNF* data were available for 17 and 39 participants in the ASD and NT groups, respectively.

In the analysis of *APOE* ε4− participants, when *Education* was added as a covariate, it was a significant predictor (*p* = 0.041), whereas T15 ∆MEP only showed a trend (*p* = 0.064), even though the model itself remained significant, *p* = 0.009, AUROC = 0.83; Dx = 83.9%. It is possible that the loss of significant effect of T15 ∆MEP was due to the listwise deletion of a few participants for whom *Education* data were not available (Table 2).

Among *APOE* ε4+ participants, T15 ∆MEP was not a significant predictor of *Diagnosis*, with or without controlling for *Education* or *Race* (*p*’s > 0.2). Choosing T30 ∆MEP as the IV and controlling for *Race* resulted in a significant model, *p* = 0.017, AUROC = 0.86; Dx = 84.2%, and both T30 ∆MEP and *Race* showed a trend-level of prediction, *p*’s < 0.1 (Table 4).

## 4. Discussion

The present study compared cTBS measures of brain plasticity in adults with HF-ASD and their age-, gender-, and IQ-matched NT controls. We found, among 63 participants including 19 participants with ASD, cTBS-induced suppression of MEPs at 15 minutes post-cTBS (T15) was a significant predictor of diagnosis with moderate (72–80%) classification accuracy, depending on the covariate included in the model. The discriminatory power of T15 ∆MEP was not driven by demographic, neuropsychological, or genetic differences between ASD and NT groups. A novel contribution of the present study was to control for influences of *BDNF* and *APOE* SNPs on cTBS aftereffects, and to assess the discriminatory power of cTBS responses between ASD and NT individuals within the more-homogenous SNP subgroups of either *BDNF* or *APOE* genes. Stratifying the study sample by either *BDNF* or *APOE* SNP status made a noticeable difference in the discriminatory power of cTBS aftereffects. While T15 ∆MEP was not a significant predictor of diagnosis among *BDNF* Met− participants, it was a stronger predictor among *BDNF* Met+ and *APOE* ε4− participants, with a classification accuracy of up to 88% and 84%, respectively. The overall pattern of cTBS aftereffects among *APOE* ε4+ was distinct from that among the other genetic subgroups, with ∆MEP at T30 showing the greatest alteration in cTBS response in ASD participants, predicting diagnosis with an accuracy of up to 84%.

### 4.1. Overall cTBS measures of plasticity in ASD and NT adults

The main finding of the present study is that, despite large inter-individual variability, M1 cTBS responses were still able to differentiate individuals with ASD from their NT counterparts in a sample that was relatively large compared to most other TBS studies (Wischnewski and Schutter 2015). The lack of a significant overall cTBS-induced suppression of MEPs in the NT group was consistent with the results of several recent studies in healthy adults (Hamada et al. 2013; Goldsworthy et al. 2014; Nettekoven et al. 2015; Vallence et al. 2015; Hordacre et al. 2017; Jannati et al. 2017, 2019). The present overall cTBS results in the NT group provide further confirmatory evidence regarding the large interindividual variability in TBS aftereffects (Corp et al. In Press).

Following previous studies investigating brain plasticity mechanisms with rTMS (Oberman et al. 2010, 2012, 2014, 2016; Fried et al. 2016, 2017; Jannati et al. 2017, 2019, 2020), we focused on the primary motor cortex in the left hemisphere, as it is typically the dominant hemisphere regardless of handedness. It is unlikely, however, that the observed differences in cTBS metrics of cortical plasticity between the two groups result from an ASD-related pathophysiological process specific to motor cortex. A structured neurological exam or medical history review found no evidence of gross or fine motor abnormalities in our ASD participants. Moreover, consistent with the results of several previous studies (Théoret et al. 2005; Minio-Paluello et al. 2009; Enticott et al. 2010, 2013b, 2013a; Oberman et al. 2012, 2016), baseline neurophysiological measures including RMT, AMT, and baseline MEP amplitude were all comparable in the two groups, confirming that baseline M1 excitability, cortico-motor reactivity, and the function of the corticospinal pathway are not affected in ASD.

The greater and longer-lasting inhibitory response to cTBS is likely due to ASD-related abnormalities in the efficacy of plasticity mechanisms in the cortex. These include both classic LTD-like synaptic plasticity as well as nonsynaptic plasticity mechanisms, including biochemical and genetic changes (Tang et al. 2017). Although abnormalities in motor domain are not among the core symptoms of ASD, motor deficits among individuals with ASD have been reported, including alterations in motor learning (Sharer et al. 2016). Moreover, there is evidence to suggest abnormalities in motor function may precede higher-level social and communication impairments, including inferring others’ intentions, gesturing, and imitation (Mostofsky and Ewen 2011; Casartelli et al. 2016). It remains unclear whether these aberrant TMS-derived physiological responses are causal or a consequence of ASD pathology. It also remains to be investigated whether and to what extent these findings translate to non-motor cortical regions. Such investigations would require combining TMS and electroencephalography (EEG) or other neuroimaging techniques (Thut et al. 2005; Thut and Pascual-Leone 2010a, 2010b; Pascual-Leone et al. 2011; Tremblay et al. 2019).

The present results confirm and expand the main finding of previous cTBS studies in adults with ASD (Oberman et al. 2012, 2016), which was abnormal plastic regulation in the form of longer-lasting cTBS aftereffects in adults with ASD compared to NT adults. Maximum sensitivity and selectivity of cTBS measures of plasticity found in the present study were comparable with those found previously, 0.75 and 0.85, respectively (Oberman et al. 2012). One noticeable difference was the substantially earlier return to baseline of cTBS-modulated MEP amplitudes in the present study (by T20) compared to those found previously, i.e., T40–T60 in the ASD group (Oberman et al. 2012). This difference in pattern of cTBS aftereffects may be attributed to interindividual variability of cTBS responses, differences in proportions of *BDNF, APOE*, and other relevant SNPs, or differences in demographics, neuropsychological characteristics, or neuroactive medications in the ASD group across studies, as suggested recently (Jannati et al. 2020).

The present results can also be considered in the context of studies on aberrant TBS responses in children and adolescents with HF-ASD (Oberman et al. 2014; Jannati et al. 2020). Those studies showed a paradoxical, facilitatory response to cTBS in approximately one-third of children and adolescents with HF-ASD (Oberman et al. 2014) or at the group level relative to typically developing children (Jannati et al. 2020). Moreover, the extent (Oberman et al. 2014) or the maximum inhibitory response to cTBS (Jannati et al. 2020) showed a maturational trajectory, increasing linearly up to the age of 16. These findings considered together with the greater and/or longer-lasting cTBS aftereffects observed in ASD adults (Oberman et al. 2012, 2016; present study) suggest a gradual change in the pattern of aberration in cTBS-derived plasticity measures in ASD across the lifespan. Individuals with ASD are more likely to show a facilitatory response to cTBS in childhood and early adolescence. As ASD individuals grow older they become more likely to show inhibitory response, with a greater inhibitory response in fully adults with ASD.

Two points are worth mentioning in comparing the present results to our recent cTBS results among healthy adults (Jannati et al. 2017). First, in that study (which included 21 of the 44 NT participants in the present study), we found T10 and T40 were the time points with the greatest explanatory power of interindividual variability in cTBS responses in healthy adults, whereas in the present study we found T15 to be the strongest predictor of diagnosis, with little discriminability between ASD and NT groups at either T10 or T40 (Figures 1–5). These contrasting results indicate that not only the most suitable post-TBS time point(s) to differentiate a given clinical population such as those with ASD from healthy individuals can differ from those in other clinical populations (e.g., McClintock et al. 2011; Tremblay et al. 2015; Fried et al. 2016), they can also differ from the time points that best characterize the gamut of TBS responses in healthy adults (Jannati et al. 2017) and across the adulthood (Freitas et al. 2011).

Second, we had found AMT was a significant predictor of cTBS response in healthy adults (Jannati et al. 2017), whereas in the present study neither AMT nor RMT or baseline MEP amplitude were significant predictors of ASD diagnosis. These results suggest, while baseline neurophysiological measures can influence TBS responses among healthy individuals (Corp et al. In Press), they may not necessarily be a good differentiator of ASD population from their healthy counterparts.

### 4.2. The roles of BDNF and APOE polymorphisms in cTBS aftereffects

Some of the factors contributing to inter-and intra-individual variability in TBS responses include activated intracortical networks (Hamada et al. 2013), functional connectivity in the motor system (Nettekoven et al. 2014, 2015), state-dependent factors (Suppa et al. 2016), and SNPs that influence neuroplasticity, including *BDNF* (Cheeran et al. 2008; Antal et al. 2010; Lee et al. 2013; Chang et al. 2014; Di Lazzaro et al. 2015; Fried et al. 2017; Jannati et al. 2017, 2019) and *APOE* (Mahley and Rall Jr 2000; White et al. 2001; Nichol et al. 2009; Wolk et al. 2010; Peña-Gomez et al. 2012; Jannati et al. 2019).

The present results on the influence of *BDNF* and *APOE* SNPs on cTBS aftereffects have two main implications: (1) The existence and pattern of abnormality in cTBS responses in ASD adults relative to NT adults are substantially modulated by both *BDNF* and *APOE* SNPs (Figures 2–5); (2) Limiting the analyses to each SNP subgroup of *BDNF* or *APOE* noticeably improves the predictive power of cTBS aftereffects in most, but not all, SNP subgroups. These two findings suggest it may not be optimal to use the same logistic-regression model for differentiating all individuals with ASD from their NT counterparts. Instead, it may be justified to devise a hierarchical decision-tree based on *BDNF* and *APOE* SNPs and perhaps other characteristics of a given individual with ASD (Tables 3 and 4). For example, if an individual with ASD is *BDNF* Met-, the next logical step may be to compare his/her cTBS response with the cTBS responses of NT individuals who have a similar *APOE* ε4 status, while also controlling for potentially important covariates. The post-cTBS time-point of interest in each step of analysis and expectations regarding the PPV and NPV of the cTBS biomarker may need to be adjusted accordingly, based on the specific SNP subgroup to which that individual belongs and perhaps demographic covariates such as race or level of education (Tables 3 and 4).

### 4.3. Additional considerations

A number of factors may limit the generalizability of the present findings. First, because our ASD sample was relatively small (n=19), it is likely there were heterogeneities among individuals with HF-ASD in demographics, clinical endophenotype, symptom severity, behavioral interventions and neuroactive medications, as well as underlying structural and functional brain differences that were not adequately captured in our sample. Such sampling errors could have reduced the representativeness of cTBS response in our ASD sample. Overcoming these limitations requires large, preferably multi-site, recruitment of individuals with ASD in future rTMS studies. Such findings would allow for better assessment of the biomarker utility of cTBS, as well as enabling more-robust indices of target engagement and therapeutic response to experimental pharmacotherapy (e.g., Lemonnier et al. 2017) and potential rTMS treatments for ASD (Cole et al. 2019). Larger samples may also allow for evaluating multivariate models that take into account several SNPs at the same time, together with other potentially important covariates, e.g., demographic, neuropsychological, and baseline neurophysiological measures, enabling a more-encompassing predictive model for the majority of individuals with ASD.

Second, in addition to the sample size, the representativeness of our ASD sample was limited in two ways: (1) because of lack of established feasibility and tolerability of rTMS in low-functioning individuals with ASD, all of our ASD participants were high-functioning; (2) because of the potential risk of rTMS-induced seizure, however small (Lerner et al. 2019), we excluded ASD participants with a history of epilepsy. The prevalence of a history of epilepsy in ASD can be as high as 26% (Viscidi et al. 2013). Moreover, a history of epilepsy is associated with more severe ASD symptoms, a history of developmental regression, and poorer adaptive and language functioning (Viscidi et al. 2013). Thus, it is possible that if our ASD group included low-functioning individuals and/or those with a history of epilepsy, the overall pattern of cTBS responses in the ASD group would be substantially different from present results. One option to study TMS measures of plasticity in individuals with ASD and current or history of epilepsy, while further reducing the potential risk of TMS-induced seizure, is to use TMS protocols such as paired associative stimulation (PAS) (Stefan et al. 2000; Ziemann 2004; see Suppa et al. 2017 for a recent review) that involve applying spTMS to the brain but still enable measuring TMS indices of plasticity in ASD (Jung et al. 2013).

Third, we were only able to stratify our sample by either *BDNF* or *APOE* SNP, as our sample size did not allow for robust assessment of the biomarker utility of cTBS aftereffects separately for each of the four present SNP subgroups (*BDNF* Met −/+ *and APOE* ε4−/+). Due to potential interactions between the effects of *BDNF* and *APOE* SNPs on rTMS plasticity metrics, the discriminatory power of cTBS aftereffects may differ substantially among those four SNP subgroups, even though, to our knowledge, such interactions have not been reported.

Lastly, another limitation of our study was the lack of data on other SNPs that influence rTMS measures of brain plasticity, e.g., the catechol-O-methyltransferase (*COMT*) Val158Met SNP (Lee et al. 2014) that can interact with the effect of *BDNF* polymorphism on rTMS responses (Witte et al. 2012).

### 4.4. Conclusions

The results of the present study show the utility of cTBS measures of M1 plasticity as a biomarker for adults with ASD, and the importance of controlling for *BDNF* and *APOE* SNPs in comparing those measures between ASD and NT individuals. Larger studies with adequate sample sizes in genetic subgroups of *BDNF, APOE*, and perhaps other potentially influential SNPs are needed to improve the assessment of the biomarker utility of TBS measures of plasticity for individuals with ASD.

## Data Availability

The datasets generated for this study are available on request to
the corresponding author.

## Acknowledgement

We thank Stephanie Changeau, Aaron Boes, and Simon Laganiere (BIDMC) for assistance with neurological examinations, and Ann Connor and Joanna Macone (BIDMC) for assistance with regulatory oversight and compliance, and evaluation of participants’ health and medical history.

This study was primarily funded by the National Institutes of Health (NIH R01 MH100186). A.P.-L. was further supported by the Sidney R. Baer Jr. Foundation, the NIH (R01 HD069776, R01 NS073601, R21 MH099196, R21 NS085491, R21 HD07616), and Harvard Catalyst | The Harvard Clinical and Translational Science Center (NCRR and the NCATS NIH, UL1 RR025758). A.J. was further supported by postdoctoral fellowships from the Natural Sciences and Engineering Research Council of Canada (NSERC 454617) and the Canadian Institutes of Health Research (CIHR 41791). L.O. was further supported by the Simons Foundation Autism Research Initiative (SFARI) and the Nancy Lurie Marks Family Foundation. A.R. was further supported by the NIH (R01 NS088583), The Boston Children’s Hospital Translational Research Program, Autism Speaks, Massachusetts Life Sciences, The Assimon Family, Brainsway, CRE Medical, Eisai, Neuroelectrics, Roche, Sage Therapeutics, and Takeda Medical. The content is solely the responsibility of the authors and does not necessarily represent the official views of Harvard, Catalyst, Harvard University and its affiliated academic health care centers, the National Institutes of Health, or any of the other listed granting agencies.

